# Clinical evaluation of BCL-2/X_L_ levels pre- and post- HER2-targeted therapy

**DOI:** 10.1101/2021.01.26.21250250

**Authors:** Jason J. Zoeller, Michael F. Press, Laura M. Selfors, Judy Dering, Dennis J. Slamon, Sara A. Hurvitz, Joan S. Brugge

## Abstract

Our previous pre-clinical work defined BCL-2 induction as a critical component of the adaptive response to lapatinib-mediated inhibition of HER2. To determine whether a similar BCL-2 upregulation occurs in lapatinib-treated patients, we evaluated gene expression within tumor biopsies, collected before and after lapatinib or trastuzumab treatment, from the TRIO-B-07 clinical trial (NCT#00769470). We detected BCL-2 mRNA upregulation in both HER2+/ER- as well as HER2+/ER+ patient tumors treated with lapatinib or trastuzumab. To address whether mRNA expression correlated with protein expression, we evaluated pre- and post-treatment tumors for BCL-2 via immunohistochemistry. Despite BCL-2 mRNA upregulation within HER2+/ER- tumors, BCL-2 protein levels were undetectable in most of the lapatinib- or trastuzumab-treated HER2+/ER- tumors. BCL-2 upregulation was evident within the majority of lapatinib-treated HER2+/ER+ tumors and was often coupled with increased ER expression and decreased proliferation. Comparable BCL-2 upregulation was not observed within the trastuzumab-treated HER2+/ER+ tumors. Together, these results provide clinical validation of the BCL-2 induction associated with the adaptive response to lapatinib and support evaluation of BCL-2 inhibitors within the context of lapatinib and other HER2-targeted receptor tyrosine kinase inhibitors.

## Introduction

Adaptive responses to targeted-therapies compromise treatment effectiveness and confer drug resistant phenotypes (1). Treatment-associated adaptive reprograming often includes parallel upregulation of alternative receptors and/or activation of alternative pathways, which compensate for target inhibition and maintain tumor growth and survival (2-8). Our previous work identified the upregulation of an anti-apoptotic program as an adaptive response to lapatinib-mediated inhibition of HER2 in vivo (9) and BEZ235-mediated inhibition of PI3K/mTOR in vitro (10). BCL-2 induction, mediated via FOXO-dependent transcription and cap-independent translation, was a critical component of this pro-survival response. To determine whether a similar anti-apoptotic program could be identified in patients treated with targeted-therapies, we utilized tumor samples that were collected during the TRIO-B-07 clinical trial (NCT#00769470). As previously described (11), TRIO-B-07 was designed to include patients with HER2+ breast cancers and biopsies before and after short-term HER2-targeted treatment with either lapatinib and/or trastuzumab.

## METHODS

### Biospecimen collection

Tumor biopsies were collected from patients enrolled in the TRIO-B-07 clinical trial (NCT#00769470) as previously described (11). TRIO-B-07 was reviewed and approved by multiple institutional review boards (UCLA; Olive View; Western), and all participants signed an IRB-approved informed consent form. The Institutional Review Board (IRB) of the Harvard University Faculty of Medicine reviewed and determined that our study is not human subjects research. Our UCLA Data Use Agreement (DUA) confirmed that all samples were de-identified and that access to identifiers (or links to identities) of individuals from whom the samples were collected would not be granted under any circumstances.

### RNA isolation and microarrays

RNA isolation and gene expression profiling (GSE130788) was performed as previously described (11). Pre-treatment and post-treatment samples from the lapatinib and trastuzumab-treatment arms of GSE130788 were subjected to Significance Analysis of Microarrays (SAM) analyses using the siggenes (v1.62.0) package in R (v4.0.1). The log10 ratios from GEO for all probes of human BCL-2 homologous and BH3-containing proteins from the BCL-2 Database (12) were tested using the single class (for post-treatment comparisons) or 2-class (for ER+/ER-pre-treatment) d.stat method. For post-treatment, delta was 0.10, corresponding to an FDR of 3.8%. For pre-treatment, delta was 1.0, corresponding to an FDR of 2.6%. Heatmaps were generated with ComplexHeatmap (v2.4.3). For genes with multiple significant probes, the probe with highest variance is presented.

### Tumor histology & IHC assays

Tissue was processed for paraffin embedding, sectioning and hematoxylin-and-eosin (H&E) staining at UCLA. Unstained sections were analyzed by IHC analysis according to previously described procedures (9). Marker specifics are as follows: BCL-2 (DAKO M0887), BCL-X_L_ (CST 2764), ER (DAKO M6364), Ki67 (DAKO M7240) and HER2 (Epitomics 42011 or DAKO A0485). BCL-2, BCL-X_L_, ER and Ki67 results were scored blindly by a board-certified pathologist using H-scores as previously described (13). For Ki67, the proportion positive reflects the total percentage of tumor cells qualitatively scored with intensities as strong (3+), moderate (2+) and weak (1+).

### Microscopy

IHC images were digitized at the DF/HCC Tissue Microarray Core or the UCLA Translational Pathology Core on the Aperio Digital Pathology Slide Scanner (Leica Biosystems). The Aperio ImageScope software (Leica Biosystems) was used for image visualization and acquisition.

### Statistics

Statistical analyses were performed using GraphPad Prism version 8 for MAC. For statistical tests, Wilcoxon matched-pairs signed rank test or Mann-Whitney test was applied. For correlation analyses, Spearman correlation was applied.

## RESULTS

To determine whether BCL-2 upregulation could be identified in patients treated with HER2-targeted therapies, we utilized tumor samples that were previously collected during the TRIO-B-07 clinical trial (NCT#00769470). TRIO-B-07 was designed to include patients with HER2+/ER- or HER2+/ER+ breast cancers and biopsies before and after 2-3 weeks of either lapatinib and/or trastuzumab treatment (**Figure 1**). Prior to definitive surgery, patients were treated with additional neo-adjuvant DNA-damaging and anti-mitotic chemotherapies. To specifically address HER2-associated events, we focused on the pre- (baseline) and post-treatment (run-in) biopsies prior to chemotherapy-treatment. To specifically compare the effects of lapatinib versus trastuzumab, we excluded tumors treated with both agents and focused on tumors treated with single agents. As previously described (11), these tumor biopsies were profiled via microarrays (GSE130788) and prepared as FFPE slides.

**Figure 1.**
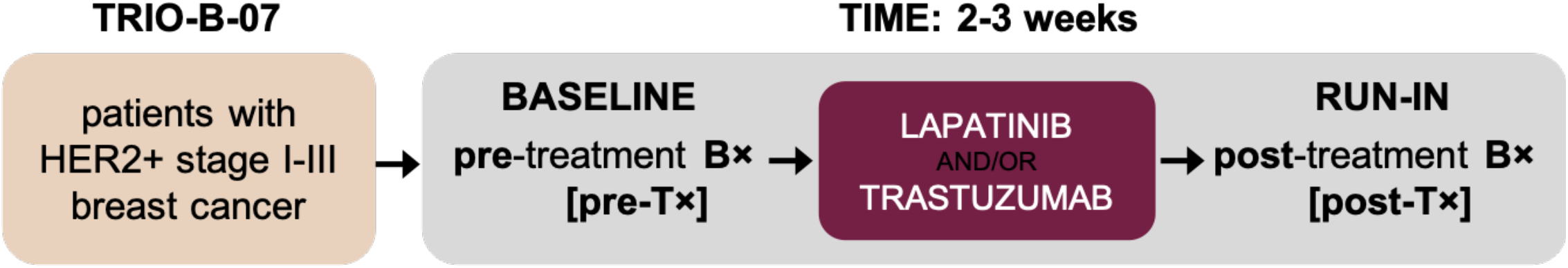
TRIO-B-07 neo-adjuvant HER2-targeted treatment course. Patients with HER2+ breast cancer underwent biopsies (B×) before (baseline; pre-treatment; pre-T×) and after (run-in; post-treatment; post-T×) two to three weeks of HER2-targeted therapies (lapatinib; L and/or trastuzumab; T). The number of paired clinical samples available for RNA or protein analysis are described within **Table 1**.

Gene expression profiles corresponding to twenty-five lapatinib-treated and twenty-six trastuzumab-treated cases were available for analysis (**Table 1**). We sub-divided the HER2+/ER- (pre-treatment ER H = 0) and HER2+/ER+ (pre-treatment ER H > 0) tumors, compared pre-treatment BCL2 mRNA levels within both sub-types and determined that HER2+/ER-tumors were associated with significantly lower BCL2 expression levels (**Supplemental Figure 1**).

**Table 1.** Patient pre- and post-treatment biopsies available for RNA or protein analysis

| **pre-Tx + post-L** | **HER2+/ER-** | **HER2+/ER+** |
| --- | --- | --- |
| RNA | 16 | 9 |
| PROTEIN | 12 | 11 |
| **pre-Tx + post-T** | **HER2+/ER-** | **HER2+/ER+** |
| RNA | 13 | 13 |
| PROTEIN | 10 | 10 |
| **NOTE** | ER- IF ER H-score = 0 | ER+ IF ER H-score > 0 |

We evaluated 2-color Agilent array data comparing the tumor biopsies collected after 2-3 weeks of treatment with either lapatinib or trastuzumab to the pre-treatment tumor biopsies from the same patient and specifically investigated the mRNA expression of BCL2 and other apoptotic regulatory genes from the BCL-2 Database (12). While there was significant alteration of several apoptotic genes within post-treatment samples, BCL2 mRNA was upregulated in HER2+/ER-(10 out of 16) as well as HER2+/ER+ (6 out of 9) lapatinib-treated tumors (**Figure 2**). BCL2 mRNA upregulation was also detected in both HER2+/ER-(9 out of 13) and HER2+/ER+ (11 out of 13) trastuzumab-treated tumors (**Figure 2**). These results indicate that BCL2 mRNA upregulation occurred in both HER2+/ER- and HER2+/ER+ patient tumors treated with lapatinib or trastuzumab.

**Figure 2.**
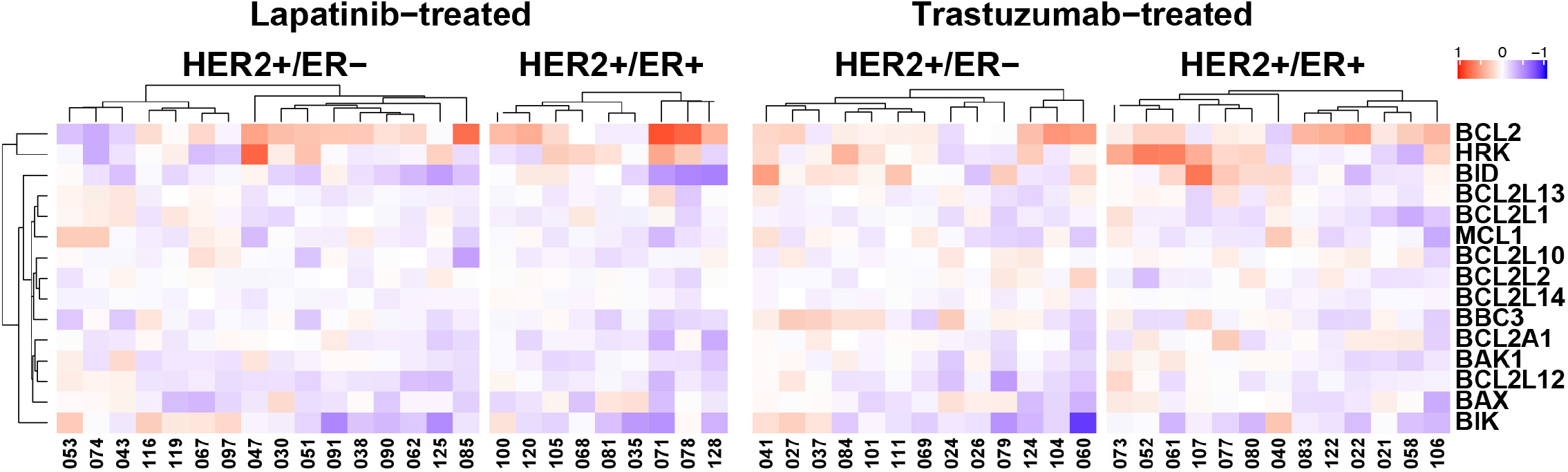
RNA-based analysis of BCL2 and other apoptotic regulatory genes. Heatmaps represent lapatinib-treated HER2+/ER-(n = 16) and HER2+/ER+ (n = 9) tumors, and trastuzumab-treated HER2+/ER-(n = 13) and HER2+/ER+ (n = 13) tumors. Each numerical identifier represents a clinical sample pair. Data are the log10 ratio of run-in (post-treatment) sample compared to paired baseline (pre-treatment) sample. Heatmaps include genes with statistically significant differences post-treatment. Heatmaps were generated with ComplexHeatmap (v2.4.3). Note, genes (proteins): BCL2 (BCL-2); HRK (HRK); BID (BID); BCL2L13 (BCL-rambo, MIL1); BCL2L1 (BCL-X_L_); MCL1 (MCL-1); BCL2L10 (NRH, BCL-B); BCL2L2 (BCL-w); BCL2L14 (BCL-G); BBC3 (PUMA); BCL2A1 (A1); BAK1 (BAK); BCL2L12 (BPR); BAX (BAX); BIK (BIK, NBK).

To address whether BCL2 mRNA expression correlated with BCL-2 protein expression, pre- and post-treatment FFPE tumors were evaluated for BCL-2 by IHC. We evaluated BCL-2 by intensity and proportion scores to calculate an H-score (13) on a case-by-case basis. Twenty-three lapatinib-treated and twenty trastuzumab-treated cases were evaluated by blinded pathological assessment (**Table 1**). In contrast to HER2+/ER+ tumors, pre-treatment BCL-2 protein levels were undetectable in most of the HER2+/ER-tumors (**Supplemental Figure 1**). Despite BCL-2 mRNA upregulation within HER2+/ER-tumors, only 2 out of 12 lapatinib-treated cases (**Figure 3a**) and 1 out of 10 trastuzumab-treated cases (**Figure 3d**) displayed a minor increase in BCL-2 protein levels following treatment. Eleven matched lapatinib-treated cases (**Figure 4a**) and ten matched trastuzumab-treated cases (**Figure 4e**) were HER2+/ER+; all of these cases were BCL-2-positive before treatment (H-scores = 15-218). Post-lapatinib, significant BCL-2 upregulation was evident in 9 out of 11 HER2+/ER+ cases (**Figure 4a**). Examples of BCL-2 and HER2 immunostaining of pre- and or post-treatment tumors from three lapatinib-treated cases [#35; #54; #71] are presented in **Figure 5**. Post-trastuzumab, BCL-2 upregulation was observed in 6 out of 10 HER2+/ER+ cases; however, these pairwise comparisons were statistically insignificant (**Figure 4e**). To determine whether treatment altered the expression of BCL-X_L_, another pro-survival protein, we performed BCL-X_L_ IHC on the lapatinib-treated and trastuzumab-treated HER2+/ER+ cases. BCL-X_L_ was upregulated in 4 out of the 9 cases where BCL-2 was upregulated in response to lapatinib (**Figure 4b**). Interestingly, upregulation in the absence of BCL-2 upregulation was detected within the remaining 2 out of 11 lapatinib-treated HER2+/ER+ tumors. **Supplemental Figure 2** includes BCL-X_L_ and BCL-2 correlation analysis for the 11 lapatinib-treated HER2+/ER+ tumors. Changes in BCL-2 and BCL-X_L_ expression were anti-correlated (r = −0.424); however, the correlation was not significant (p = 0.192). For the trastuzumab-treated HER2+/ER+ cases, BCL-X_L_ was upregulated in only 1 out of the 6 cases where BCL-2 was upregulated (**Figure 4f**) and within two additional cases; however, one of these cases displayed only a minor increase in BCL-X_L_ protein levels post-treatment. These results indicate that BCL-2 is upregulated within the majority of lapatinib-treated HER2+/ER+ tumors and that BCL-2 protein is undetectable pre- and post-treatment in most HER2+/ER-tumors.

**Figure 3.**
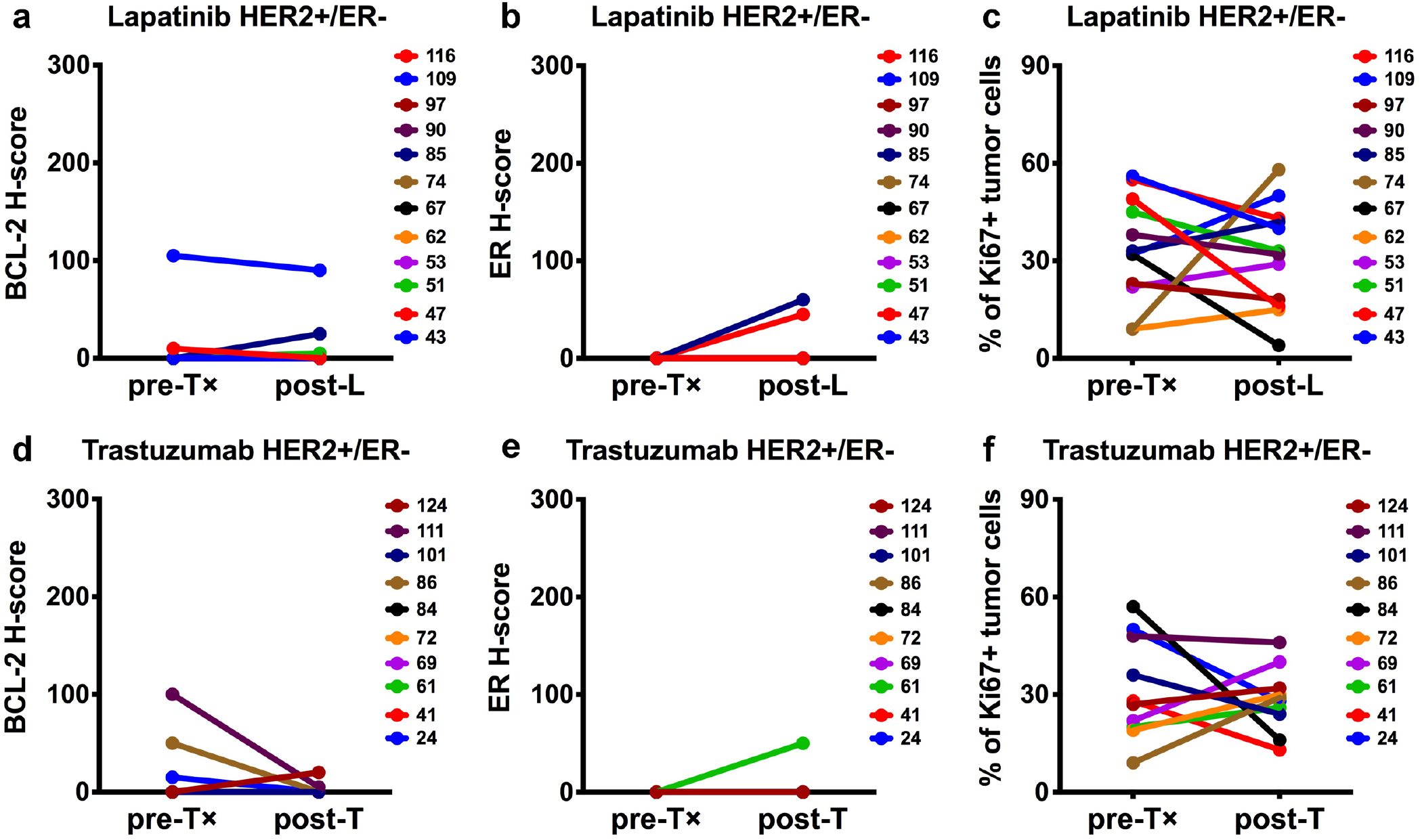
[HER2+/ER-] Comparison of BCL-2, ER and Ki67 protein levels pre-and post-lapatinib [L] or post-trastuzumab [T] treatment. Twelve HER2+/ER-lapatinib-treated (**a**-**c**) and ten HER2+/ER-trastuzumab-treated (**d**-**f**) sample pairs were evaluated for BCL-2 (**a** and **d**), ER (**b** and **e**) and Ki67 (**c** and **f**). BCL-2 and ER were semi-quantitated via H-score assessment. Ki67 values represent the estimated percentage of Ki67+ tumor cells. Each numerical identifier represents a clinical sample pair. Each plot compares baseline (pre-treatment) and run-in (post-treatment) paired samples. Statistical comparisons utilized Wilcoxon matched-pairs signed rank two-tailed test (p-values > 0.05). p-values = [**a**] >0.9999; [**b**] 0.5000; [**c**] 0.6353; [**d**] 0.3750; [**e**] >0.9999; [**f**] 0.7695.

**Figure 4.**
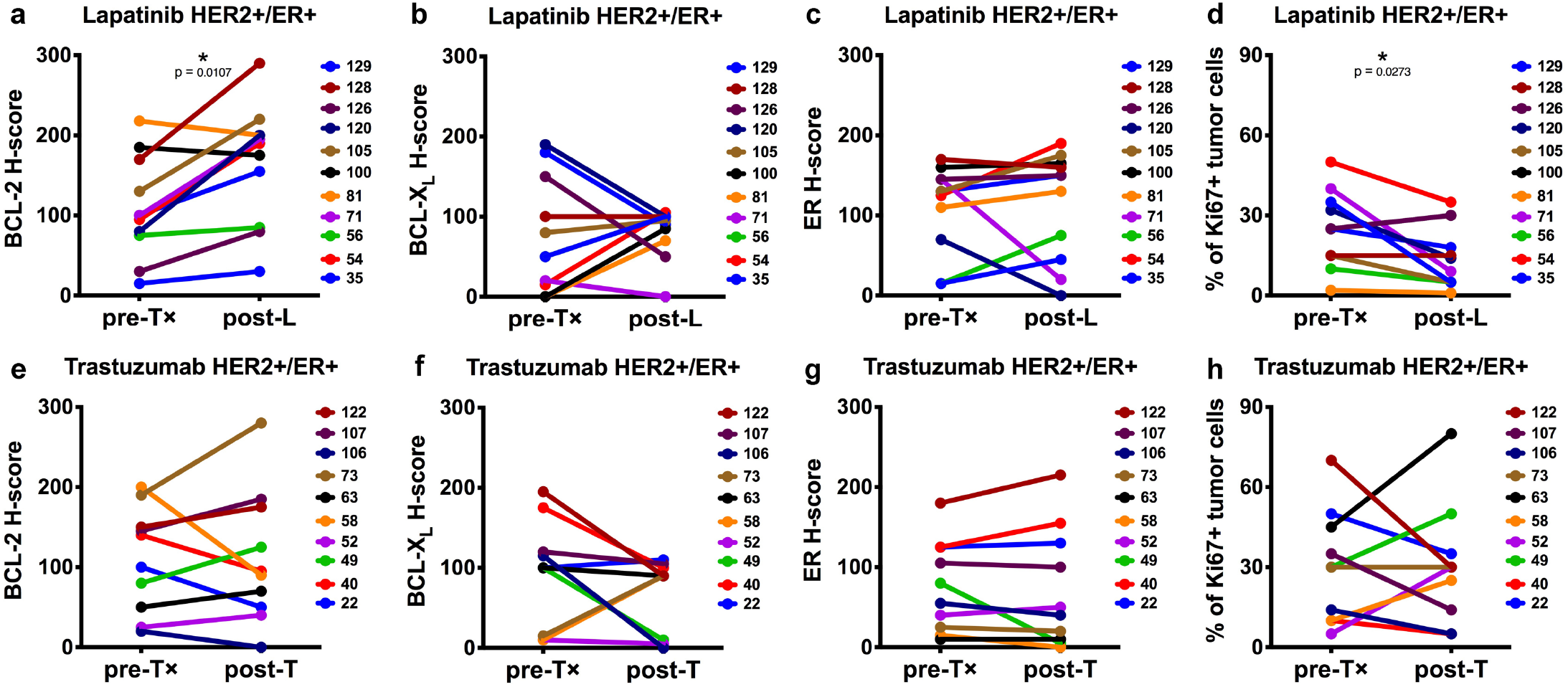
[HER2+/ER+] Comparison of BCL-2, BCL-X_L_, ER and Ki67 protein levels pre- and post-lapatinib [L] or post-trastuzumab [T] treatment. Eleven HER2+/ER+ lapatinib-treated (**a**-**d**) and ten HER2+/ER+ trastuzumab-treated (**e**-**h**) sample pairs were evaluated for BCL-2 (**a** and **e**), BCL-X_L_ (**b** and **f**), ER (**c** and **g**) and Ki67 (**d** and **h**). BCL-2, BCL-X_L_, ER and Ki67 were evaluated as described in **Figure 3**. Each numerical identifier represents a clinical sample pair. Each plot compares baseline (pre-treatment) and run-in (post-treatment) paired samples. Statistical comparisons utilized Wilcoxon matched-pairs signed rank two-tailed test and p-values < 0.05 are indicated (*). p-values = [**a**] 0.0107; [**b**] 0.9727; [**c**] 0.4521; [**d**] 0.0273; [**e**] 0.9922; [**f**] 0.2207; [**g**] 0.8945; [**h**] 0.9297.

**Figure 5.**
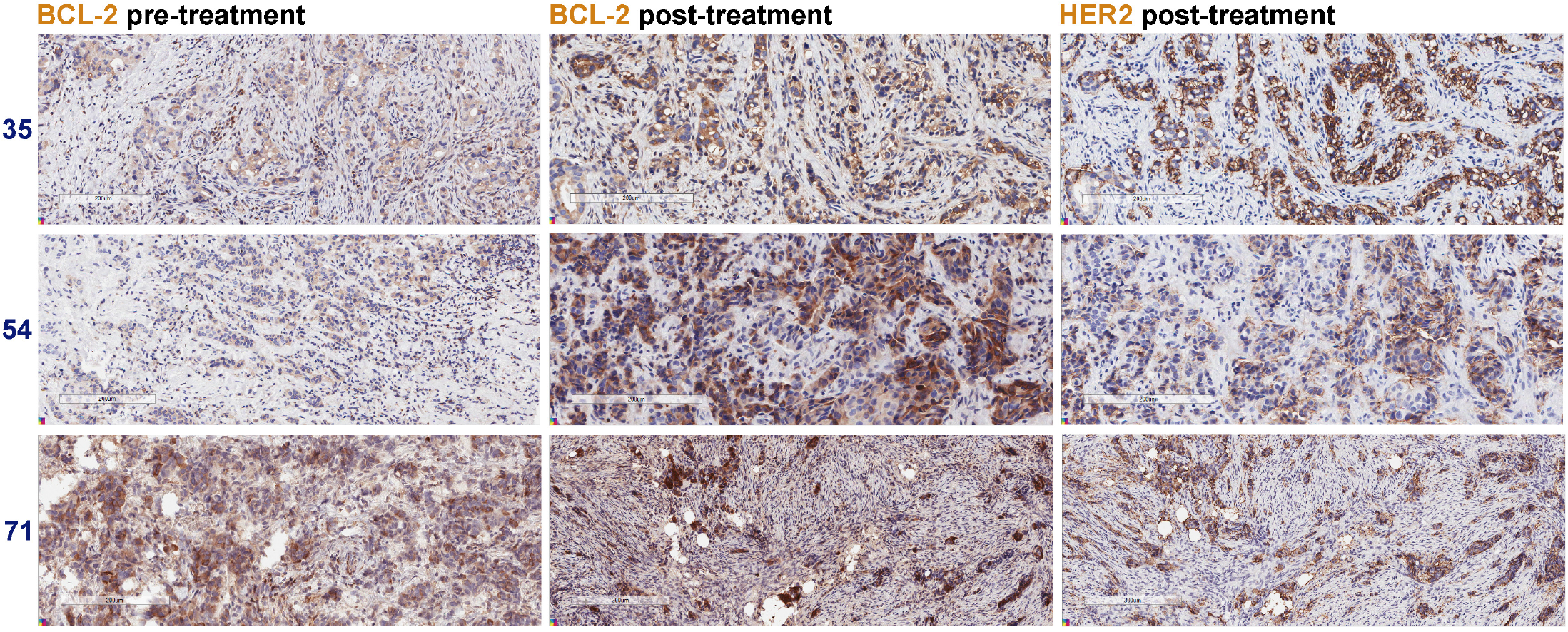
Lapatinib-induced BCL-2 upregulation post-treatment. Comparison of BCL-2 levels pre- and post-treatment for cases #35, #54 and #71. HER2 serial section IHC identified the tumor cells post-lapatinib. Scale bars, 200-300 μm.

Since BCL2 is a direct transcriptional target of ER (14), we performed ER IHC and examined whether lapatinib-associated BCL-2 induction paralleled ER upregulation. ER was upregulated in 6 out of the 9 HER2+/ER+ cases where BCL-2 was upregulated in response to lapatinib (**Figure 4c**) and in 2 out of the 6 HER2+/ER+ cases where BCL-2 was upregulated in response to trastuzumab (**Figure 4g** and **Supplemental Figure 2**). By comparison, ER upregulation was detected within 2 out of 12 HER2+/ER-lapatinib-treated cases (**Figure 3b**) and 1 out of 10 HER2+/ER-trastuzumab-treated cases (**Figure 3e**); however, only one of these cases displayed a minor increase in BCL-2. These results indicate that BCL-2 upregulation in lapatinib-treated HER2+/ER+ tumors is often, but not always, coupled with increased ER expression.

To determine whether BCL-2 upregulation correlated with the inhibition of tumor proliferation, we performed Ki67 IHC. Pair-wise comparisons revealed non-significant Ki67 alterations within the HER2+/ER+ tumors post-trastuzumab (**Figure 4h**) and the HER2+/ER-tumors treated with either lapatinib (**Figure 3c**) or trastuzumab (**Figure 3f**). For HER2+/ER+ tumors, we detected an overall reduction in Ki67+ tumor cells post-lapatinib (**Figure 4d**). The percentage of Ki67+ tumor cells was reduced in the majority of cases where BCL-2 was upregulated in response to lapatinib. Correlation analysis (**Supplemental Figure 2**) supported these observations and indicated BCL-2 and Ki67 were anti-correlated (r = −0.421); however, the correlation was not significant (p = 0.196). These results indicate that BCL-2 upregulation within lapatinib-treated HER2+/ER+ tumors is frequently associated with lapatinib-induced proliferative blockade.

## DISCUSSION

Here, we provide clinical validation of the BCL-2 induction associated with the adaptive response to lapatinib-mediated HER2 blockade. The upregulation of BCL2 mRNA occurred in both HER2+/ER- as well as HER2+/ER+ patient tumors treated with lapatinib or trastuzumab; however, BCL-2 protein levels pre- and post-treatment were undetectable in most HER2+/ER-tumors. The upregulation of BCL-2 protein was evident within the majority of lapatinib-treated HER2+/ER+ tumors and was often coupled with increased ER expression and decreased proliferation. Comparable BCL-2 upregulation was not observed within the trastuzumab-treated HER2+/ER+ tumors. Together, these results support clinical evaluation of BCL-2 inhibitors within the context of lapatinib and the treatment of HER2+/ER+ breast cancers.

Our results are consistent with in vitro studies of ER-mediated lapatinib resistance mechanisms (15, 16), which provided evidence for ER and BCL-2 induction within lapatinib-resistant models. Our results are further supported by similar lapatinib-focused analyses of ER and BCL-2 co-expression (17), which demonstrated parallel upregulation of ER and BCL-2 within two pre-clinical models of treatment resistant HER2+ breast cancer as well as a subset of lapatinib-treated HER2+ clinical samples. The relationship between lapatinib-mediated HER2 blockade and ER induction is consistent with evidence that pathways activated (MAPK/ERK or PI3K/AKT) downstream of HER2-signaling negatively regulate ER through both epigenetic and transcriptional mechanisms (15, 18-24); whereas, the correlation between BCL-2 upregulation and ER upregulation is consistent with evidence that BCL-2 is a direct target of ER transcriptional regulation (14). Since BCL-2 upregulation was also observed in a few tumors without ER upregulation, our results also suggest that there can be ER-independent regulation of BCL-2. These results are consistent with multiple previous reports demonstrating that anti-estrogen treatment only decreased BCL-2 expression in a subset of tumors analyzed (25-28).

The correlation between BCL-2 upregulation and decreased proliferation is consistent with evidence that HER2 inhibition confers adaptive reprogramming. Since lapatinib and trastuzumab have differential mechanisms of action (29) and variable impacts on HER2-signaling blockade (30), the lapatinib-associated BCL-2 responses could be directly related to superior HER2 inhibition achieved with lapatinib versus trastuzumab.

Endpoint analysis of pathological responses in the TRIO-B-07 clinical trial indicated that lapatinib-treated HER2+/ER+ tumors had the lowest pathological complete response rates (11). Since these tumors were subsequently treated with additional neo-adjuvant DNA-damaging and anti-mitotic chemotherapies, we are unable to address the extent to which BCL-2 induction clinically impacted the lapatinib-associated pathological responses observed within HER2+/ER+ tumors. Nonetheless, we would predict that tumor populations distinguished by elevated BCL-2 could contribute to residual and recurrent disease. These predictions are based upon our own pre-clinical in vivo data demonstrating BCL-2 induction in lapatinib-treated HER2+ residual tumors (9) as well as additional in vitro and in vivo studies demonstrating BCL-2 induction within lapatinib-resistant models (15-17).

BCL-2 dependencies have been described with the context of HER2-/ER+ breast cancers, which are enriched for BCL-2 expression (31-33). Based upon these observations, pre-clinical studies have determined that selective targeting of BCL-2 via ABT-199/venetoclax (34) enhanced the effectiveness of ER-targeted treatment (35) and have motivated clinical investigations of venetoclax combined treatment strategies within HER2-/ER+ tumors (36). Since our results demonstrated similar BCL-2 enrichment within the context of HER2+/ER+ breast cancers, we would predict that HER2-targeted and ER-targeted treatments of these tumor types could be enhanced via venetoclax.

The pro-survival functions of BCL-2, BCL-X_L_ and MCL-1 have been described within various contexts of HER2-targeted therapies. Using cell-derived and patient-derived xenograft models of HER2+/ER-breast tumors, our previous pre-clinical work provides evidence that dual targeting of BCL-2 and BCL-X_L_, via ABT-737 (37) or ABT-263/navitoclax (38), enhanced the in vivo effectiveness of lapatinib (9) or T-DM1 (39). Another report identified an increased BCL-2:BAX ratio within a trastuzumab-resistant in vitro model and demonstrated that dual targeting of BCL-2 and BCL-X_L_ enhanced the effectiveness of trastuzumab (40). Multiple pre-clinical studies have identified MCL-1 as a critical component of lapatinib-resistant phenotypes and demonstrated that blockade of MCL-1, via obatoclax (41) or S63845 (42), enhanced treatment effectiveness (43-47). Although several studies have demonstrated trastuzumab-associated downregulation of MCL-1 (48) or BCL-2 (49), these studies also reported that further disruption or modulation of the pro-survival proteins enhanced treatment effectiveness. Together with our TRIO-B-07 clinical results, these studies support the evaluation of BCL-targeted agents within the context of HER2-targeted treatments.

## Data Availability

The TRIO-B-07 data files are publicly available from the GEO database (GSE130788).

## ACKNOWLEDGEMENTS

We would like to thank Enrique Guandique (UCLA), Lillian Ramos (UCLA) and Aleksandr Vagodny (Harvard Medical School) for IHC-related technical assistance. We would like to acknowledge the Dana-Farber Harvard Cancer Center Tissue Microarray Imaging Core and UCLA Translational Pathology Core for access to and usage of the Aperio slide scanner.

## AUTHORS’ CONTRIBUTIONS

J.J.Z. designed and performed all IHC assays and drafted the manuscript.

M.F.P. evaluated all IHC with J.J.Z.

L.M.S. evaluated the microarray datasets.

J.D. evaluated the microarray datasets.

D.J.S. designed the TRIO-B-07 clinical trial (NCT#00769470).

S.A.H. designed and led the TRIO-B-07 clinical trial (NCT#00769470).

J.S.B. contributed to data analysis and preparation of the manuscript.

## List of Abbreviations

B×: biopsy
DUA: Data Usage Agreement
DF: Dana-Farber
GSE: Gene Series Expression
HCC: Harvard Cancer Center
HER2: human epidermal growth factor receptor-2
ER: estrogen receptor
FOXO: members of the class O of forkhead box transcription factors
H&E: hematoxylin-and-eosin
H-score: histo-score
IHC: immunohistochemistry
IRB: institutional review board
L: lapatinib
mTOR: mammalian target of rapamycin
NCT: National Clinical Trial
p: p-value
PI3K: phosphoinositide 3-kinase
r: Spearman correlation
RNA: ribonucleic acid
SAM: Significance Analysis of Microarrays
T: trastuzumab
T×: treatment
UCLA: University of California Los Angeles

---

**Supplemental Figure 1.**
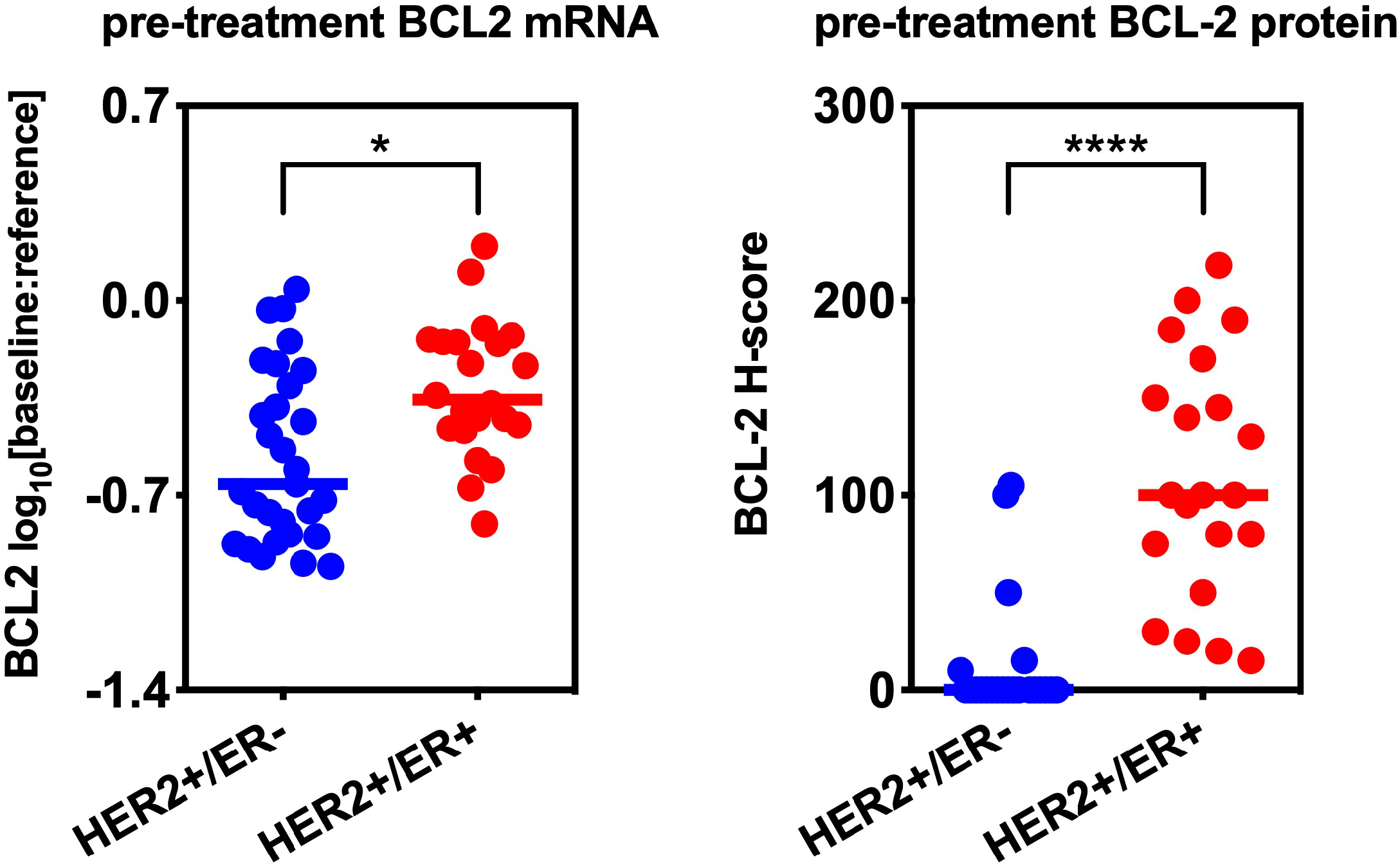
Pre-treatment expression levels of BCL2 mRNA or BCL-2 protein. Each plot compares baseline (pre-treatment) HER2+/ER-(pre-treatment ER H = 0) and HER2+/ER+ (pre-treatment ER H > 0) tumors. Statistical comparisons of BCL2 mRNA utilized Significance Analysis of Microarrays (SAM) analyses, FDR-p = 0.0178 (*). Each line represents the median. Statistical comparisons of BCL-2 protein utilized Mann-Whitney two-tailed test, p < 0.0001 (****). Each line represents the median.

**Supplemental Figure 2.**
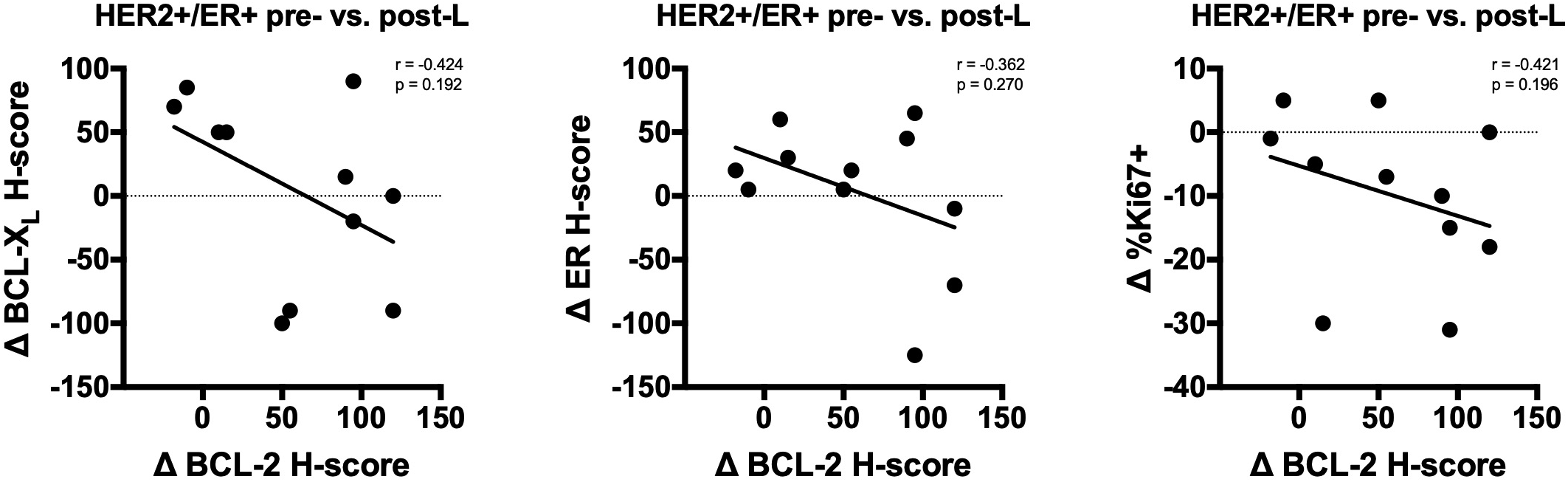
Correlation analyses for lapatinib-treated HER2+/ER+ tumors. Each graph compares BCL-2 and BCL-X_L_, ER or Ki67 alterations (Δ = **Figure 4** post-value – **Figure 4** pre-value). The Spearman correlation [r] and associated p-value [p] is indicated for each comparison.

## REFERENCES

1. Pazarentzos E, Bivona TG. Adaptive stress signaling in targeted cancer therapy resistance. Oncogene. 2015;34(45):5599–606.

2. Chandarlapaty S, Sawai A, Scaltriti M, Rodrik-Outmezguine V, Grbovic-Huezo O, Serra V, et al. AKT inhibition relieves feedback suppression of receptor tyrosine kinase expression and activity. Cancer Cell. 2011;19(1):58–71.

3. Serra V, Scaltriti M, Prudkin L, Eichhorn PJ, Ibrahim YH, Chandarlapaty S, et al. PI3K inhibition results in enhanced HER signaling and acquired ERK dependency in HER2-overexpressing breast cancer. Oncogene. 2011;30(22):2547–57.

4. Rodrik-Outmezguine VS, Chandarlapaty S, Pagano NC, Poulikakos PI, Scaltriti M, Moskatel E, et al. mTOR kinase inhibition causes feedback-dependent biphasic regulation of AKT signaling. Cancer Discov. 2011;1(3):248–59.

5. Garrett JT, Olivares MG, Rinehart C, Granja-Ingram ND, Sanchez V, Chakrabarty A, et al. Transcriptional and posttranslational up-regulation of HER3 (ErbB3) compensates for inhibition of the HER2 tyrosine kinase. Proc Natl Acad Sci U S A. 2011;108(12):5021–6.

6. Duncan JS, Whittle MC, Nakamura K, Abell AN, Midland AA, Zawistowski JS, et al. Dynamic reprogramming of the kinome in response to targeted MEK inhibition in triple-negative breast cancer. Cell. 2012;149(2):307–21.

7. Sun C, Wang L, Huang S, Heynen GJ, Prahallad A, Robert C, et al. Reversible and adaptive resistance to BRAF(V600E) inhibition in melanoma. Nature. 2014;508(7494):118–22.

8. Stuhlmiller TJ, Miller SM, Zawistowski JS, Nakamura K, Beltran AS, Duncan JS, et al. Inhibition of Lapatinib-Induced Kinome Reprogramming in ERBB2-Positive Breast Cancer by Targeting BET Family Bromodomains. Cell Rep. 2015;11(3):390–404.

9. Zoeller JJ, Bronson RT, Selfors LM, Mills GB, Brugge JS. Niche-localized tumor cells are protected from HER2-targeted therapy via upregulation of an anti-apoptotic program in vivo. npj Breast Cancer. 2017;3(18).

10. Muranen T, Selfors LM, Worster DT, Iwanicki MP, Song L, Morales FC, et al. Inhibition of PI3K/mTOR leads to adaptive resistance in matrix-attached cancer cells. Cancer Cell. 2012;21(2):227–39.

11. Hurvitz SA, Caswell-Jin JL, McNamara KL, Zoeller JJ, Bean GR, Dichmann R, et al. Pathologic and molecular responses to neoadjuvant trastuzumab and/or lapatinib from a phase II randomized trial in HER2-positive breast cancer (TRIO-US B07). Nature communications. 2020;11(1):5824.

12. Rech de Laval V, Deléage G, Aouacheria A, Combet C. BCL2DB: database of BCL-2 family members and BH3-only proteins. Database. 2014;2014.

13. Goulding H, Pinder S, Cannon P, Pearson D, Nicholson R, Snead D, et al. A new immunohistochemical antibody for the assessment of estrogen receptor status on routine formalin-fixed tissue samples. Hum Pathol. 1995;26(3):291–4.

14. Perillo B, Sasso A, Abbondanza C, Palumbo G. 17beta-estradiol inhibits apoptosis in MCF-7 cells, inducing bcl-2 expression via two estrogen-responsive elements present in the coding sequence. Mol Cell Biol. 2000;20(8):2890–901.

15. Xia W, Bacus S, Hegde P, Husain I, Strum J, Liu L, et al. A model of acquired autoresistance to a potent ErbB2 tyrosine kinase inhibitor and a therapeutic strategy to prevent its onset in breast cancer. Proc Natl Acad Sci U S A. 2006;103(20):7795–800.

16. Wang YC, Morrison G, Gillihan R, Guo J, Ward RM, Fu X, et al. Different mechanisms for resistance to trastuzumab versus lapatinib in HER2-positive breast cancers--role of estrogen receptor and HER2 reactivation. Breast Cancer Res. 2011;13(6):R121.

17. Giuliano M, Hu H, Wang YC, Fu X, Nardone A, Herrera S, et al. Upregulation of ER Signaling as an Adaptive Mechanism of Cell Survival in HER2-Positive Breast Tumors Treated with Anti-HER2 Therapy. Clin Cancer Res. 2015;21(17):3995–4003.

18. Oh AS, Lorant LA, Holloway JN, Miller DL, Kern FG, El-Ashry D. Hyperactivation of MAPK induces loss of ERalpha expression in breast cancer cells. Mol Endocrinol. 2001;15(8):1344–59.

19. Holloway JN, Murthy S, El-Ashry D. A cytoplasmic substrate of mitogen-activated protein kinase is responsible for estrogen receptor-alpha down-regulation in breast cancer cells: the role of nuclear factor-kappaB. Mol Endocrinol. 2004;18(6):1396–410.

20. Creighton CJ, Hilger AM, Murthy S, Rae JM, Chinnaiyan AM, El-Ashry D. Activation of mitogen-activated protein kinase in estrogen receptor alpha-positive breast cancer cells in vitro induces an in vivo molecular phenotype of estrogen receptor alpha-negative human breast tumors. Cancer Res. 2006;66(7):3903–11.

21. Bayliss J, Hilger A, Vishnu P, Diehl K, El-Ashry D. Reversal of the estrogen receptor negative phenotype in breast cancer and restoration of antiestrogen response. Clin Cancer Res. 2007;13(23):7029–36.

22. Plotkin A, Volmar CH, Wahlestedt C, Ayad N, El-Ashry D. Transcriptional repression of ER through hMAPK dependent histone deacetylation by class I HDACs. Breast Cancer Res Treat. 2014;147(2):249–63.

23. Guo S, Sonenshein GE. Forkhead box transcription factor FOXO3a regulates estrogen receptor alpha expression and is repressed by the Her-2/neu/phosphatidylinositol 3-kinase/Akt signaling pathway. Mol Cell Biol. 2004;24(19):8681–90.

24. Creighton CJ, Fu X, Hennessy BT, Casa AJ, Zhang Y, Gonzalez-Angulo AM, et al. Proteomic and transcriptomic profiling reveals a link between the PI3K pathway and lower estrogen-receptor (ER) levels and activity in ER+ breast cancer. Breast Cancer Res. 2010;12(3):R40.

25. Johnston SR, MacLennan KA, Sacks NP, Salter J, Smith IE, Dowsett M. Modulation of Bcl-2 and Ki-67 expression in oestrogen receptor-positive human breast cancer by tamoxifen. European journal of cancer (Oxford, England : 1990). 1994;30a(11):1663–9.

26. Keen JC, Dixon JM, Miller EP, Cameron DA, Chetty U, Hanby A, et al. The expression of Ki-S1 and BCL-2 and the response to primary tamoxifen therapy in elderly patients with breast cancer. Breast Cancer Res Treat. 1997;44(2):123–33.

27. Cameron DA, Keen JC, Dixon JM, Bellamy C, Hanby A, Anderson TJ, et al. Effective tamoxifen therapy of breast cancer involves both antiproliferative and pro-apoptotic changes. European journal of cancer (Oxford, England : 1990). 2000;36(7):845–51.

28. Kenny FS, Willsher PC, Gee JM, Nicholson R, Pinder SE, Ellis IO, et al. Change in expression of ER, bcl-2 and MIB1 on primary tamoxifen and relation to response in ER positive breast cancer. Breast Cancer Res Treat. 2001;65(2):135–44.

29. Hurvitz SA, Hu Y, O’Brien N, Finn RS. Current approaches and future directions in the treatment of HER2- positive breast cancer. Cancer Treat Rev. 2013;39(3):219–29.

30. Diermeier-Daucher S, Breindl S, Buchholz S, Ortmann O, Brockhoff G. Modular anti-EGFR and anti-Her2 targeting of SK-BR-3 and BT474 breast cancer cell lines in the presence of ErbB receptor-specific growth factors. Cytometry Part A : the journal of the International Society for Analytical Cytology. 2011;79(9):684–93.

31. Silvestrini R, Veneroni S, Daidone MG, Benini E, Boracchi P, Mezzetti M, et al. The Bcl-2 protein: a prognostic indicator strongly related to p53 protein in lymph node-negative breast cancer patients. J Natl Cancer Inst. 1994;86(7):499–504.

32. Leek RD, Kaklamanis L, Pezzella F, Gatter KC, Harris AL. bcl-2 in normal human breast and carcinoma, association with oestrogen receptor-positive, epidermal growth factor receptor-negative tumours and in situ cancer. Br J Cancer. 1994;69(1):135–9.

33. Bhargava V, Kell DL, van de Rijn M, Warnke RA. Bcl-2 immunoreactivity in breast carcinoma correlates with hormone receptor positivity. Am J Pathol. 1994;145(3):535–40.

34. Souers AJ, Leverson JD, Boghaert ER, Ackler SL, Catron ND, Chen J, et al. ABT-199, a potent and selective BCL-2 inhibitor, achieves antitumor activity while sparing platelets. Nat Med. 2013;19(2):202–8.

35. Vaillant F, Merino D, Lee L, Breslin K, Pal B, Ritchie ME, et al. Targeting BCL-2 with the BH3 mimetic ABT- 199 in estrogen receptor-positive breast cancer. Cancer Cell. 2013;24(1):120–9.

36. Lok SW, Whittle JR, Vaillant F, Teh CE, Lo LL, Policheni AN, et al. A Phase Ib Dose-Escalation and Expansion Study of the BCL2 Inhibitor Venetoclax Combined with Tamoxifen in ER and BCL2-Positive Metastatic Breast Cancer. Cancer Discov. 2019;9(3):354–69.

37. Oltersdorf T, Elmore SW, Shoemaker AR, Armstrong RC, Augeri DJ, Belli BA, et al. An inhibitor of Bcl-2 family proteins induces regression of solid tumours. Nature. 2005;435(7042):677–81.

38. Tse C, Shoemaker AR, Adickes J, Anderson MG, Chen J, Jin S, et al. ABT-263: a potent and orally bioavailable Bcl-2 family inhibitor. Cancer Res. 2008;68(9):3421–8.

39. Zoeller JJ, Vagodny A, Taneja K, Tan BY, O’Brien N, Slamon DJ, et al. Neutralization of BCL-2/XL Enhances the Cytotoxicity of T-DM1 In Vivo. Mol Cancer Ther. 2019;18(6):1115–26.

40. Crawford A, Nahta R. Targeting Bcl-2 in Herceptin-Resistant Breast Cancer Cell Lines. Curr Pharmacogenomics Person Med. 2011;9(3):184–90.

41. Nguyen M, Marcellus RC, Roulston A, Watson M, Serfass L, Murthy Madiraju SR, et al. Small molecule obatoclax (GX15-070) antagonizes MCL-1 and overcomes MCL-1-mediated resistance to apoptosis. Proc Natl Acad Sci U S A. 2007;104(49):19512–7.

42. Kotschy A, Szlavik Z, Murray J, Davidson J, Maragno AL, Le Toumelin-Braizat G, et al. The MCL1 inhibitor S63845 is tolerable and effective in diverse cancer models. Nature. 2016;538(7626):477–82.

43. Martin AP, Miller A, Emad L, Rahmani M, Walker T, Mitchell C, et al. Lapatinib resistance in HCT116 cells is mediated by elevated MCL-1 expression and decreased BAK activation and not by ERBB receptor kinase mutation. Mol Pharmacol. 2008;74(3):807–22.

44. Martin AP, Mitchell C, Rahmani M, Nephew KP, Grant S, Dent P. Inhibition of MCL-1 enhances lapatinib toxicity and overcomes lapatinib resistance via BAK-dependent autophagy. Cancer Biol Ther. 2009;8(21):2084–96.

45. Mitchell C, Yacoub A, Hossein H, Martin AP, Bareford MD, Eulitt P, et al. Inhibition of MCL-1 in breast cancer cells promotes cell death in vitro and in vivo. Cancer Biol Ther. 2010;10(9):903–17.

46. Merino D, Whittle JR, Vaillant F, Serrano A, Gong JN, Giner G, et al. Synergistic action of the MCL-1 inhibitor S63845 with current therapies in preclinical models of triple-negative and HER2-amplified breast cancer. Sci Transl Med. 2017;9(401).

47. Eustace AJ, Conlon NT, McDermott MSJ, Browne BC, O’Leary P, Holmes FA, et al. Development of acquired resistance to lapatinib may sensitise HER2-positive breast cancer cells to apoptosis induction by obatoclax and TRAIL. BMC Cancer. 2018;18(1):965.

48. Henson ES, Hu X, Gibson SB. Herceptin sensitizes ErbB2-overexpressing cells to apoptosis by reducing antiapoptotic Mcl-1 expression. Clin Cancer Res. 2006;12(3 Pt 1):845-53.

49. Milella M, Trisciuoglio D, Bruno T, Ciuffreda L, Mottolese M, Cianciulli A, et al. Trastuzumab down-regulates Bcl-2 expression and potentiates apoptosis induction by Bcl-2/Bcl-XL bispecific antisense oligonucleotides in HER-2 gene--amplified breast cancer cells. Clin Cancer Res. 2004;10(22):7747–56.

